# Effectiveness of a Pragmatic Metabolic Care Clinic for Patients with Severe Mental Illness: Protocol for the Randomized Controlled Meta-Care Trial

**DOI:** 10.64898/2026.02.28.26347334

**Authors:** Grimur Høgnason Mohr, Sri Mahavir Agarwal, Victor Sørensen, Cecilie Koldbæk Lemvigh, Mikkel Erlang Sørensen, Marcos Sanches, Anne Regitze Hartmann Hamilton, Carlo Alberto Barcella, Dan Siskind, Julie Midtgaard, Tina Vilsbøll, Margaret K. Hahn, Bjørn H. Ebdrup

**Affiliations:** Center for Neuropsychiatric Schizophrenia Research (CNSR), Mental Health Center, Glostrup, Copenhagen University Hospital – Mental Health Services CPH, Copenhagen, Denmark; Schizophrenia Division, Centre for Addiction and Mental Health (CAMH), Toronto, Canada; Centre for Applied Research in Mental health Care, CARMEN, Copenhagen University Hospital, Mental Health Centre Glostrup, Denmark; Department of Cardiology, Copenhagen University Hospital Herlev and Gentofte, Denmark; School of Clinical Medicine, The University of Queensland, Woolloongabba, QLD, Australia; Department of Clinical Medicine, Faculty of Health and Medical Sciences, University of Copenhagen, Copenhagen, Denmark; Clinical Research, Steno Diabetes Center Copenhagen, Herlev Hospital, Denmark

**Author notes:** Corresponding author: Grimur Høgnason Mohr, contact, Trial sponsor: Bjørn H. Ebdrup, contact, Address: Nordstjernevej 41, 2600 Glostrup, Denmark. Equal first author contribution (Grimur Høgnason Mohr, Sri Mahavir Agarwal). Equal senior author contribution (Margaret K. Hahn, Bjørn H. Ebdrup).

## Abstract

**Introduction:** Severe mental illness is associated with high mortality rates and cardiovascular disease. Obesity and dysmetabolism associated with antipsychotic treatment comprise modifiable risk factors, which remain undertreated. Interventions such as antipsychotic-switching, lifestyle-interventions and weight-reducing medication have shown varying results indicating a need for a more individualized approach. The Meta-Care Trial aims to assess the effectiveness of a pragmatic, individualized, evidence- and guideline-based cardiometabolic intervention.

**Methods and analysis:** Meta-Care is an open-label randomized controlled trial (RCT). Patients between 18-45 years with schizophrenia spectrum disorders or bipolar disorder will be recruited from in- and outpatient Mental Health Services in the Capital Region of Denmark. Inclusion criteria include treatment with antipsychotics and: either i) ≥5% body weight gain or ≥5cm waist circumference increase since initiation of antipsychotic therapy, or ii) a body mass index (BMI) ≥30 kg/m^2^, or iii) BMI ≥27 kg/m^2^ and related cardiovascular risk factors. Patients are randomized to a pragmatic, individualized metabolic clinic using evidence- and guideline-based care in a mental health center or standard care. Allocation-ratio is 1:1.

The *primary outcome* is the proportion of patients achieving weight loss ≥5% of initial body weight after 12 months. *Secondary and exploratory outcomes* cover cardiometabolic risk factors, cognition, personal recovery, and quality of life. Qualitative interviews will explore patient experience and contextual factors. Recruitment started in October 2023 and will include a total of 84 patients.

**Ethics and dissemination:** The Meta-Care trial is funded by The Independent Research Fund Denmark and The Wørzner Memorial Fund for Research in Mental Illness. The trial has been approved by the Regional Ethics Committee and Data Protection Agency in the Capital Region of Denmark. Positive, negative, and inconclusive results will be published in scientific peer-reviewed journals, presented at conferences, and dispersed to patient organisations and media.

**Strengths and limitations:**

- The Meta-Care Trial is the first randomized control trial (RCT) to investigate the effectiveness and acceptability of a pragmatic, individualized metabolic clinic located in a mental health center using evidence- and guideline-based care to treat obesity and cardiometabolic risk factors in patients with severe mental illness
- The pragmatic design with limited exclusion criteria and simple outcome measures will generate results that are generalizable to clinical practice
- The complex Meta-Care multi-intervention limits inferences of effects explained by specific modifications of pharmacotherapy or lifestyle changes
- Potential knowledge exchange from treating personnel in the Meta-Care Trial to caregivers in the standard care group may lead to contamination bias
- Although the Meta-Care trial has an open label design, measurements of primary and secondary outcomes will be carried out by blinded assessors

## Introduction

Patients with severe mental illness (SMI), such as schizophrenia spectrum disorders and bipolar disorder, have a decreased life expectancy compared to the general population and mortality rates have not improved during the past decades^1,2^. The reduced life expectancy is largely explained by alarmingly high rates of somatic multimorbidity and a lifetime suicide rate of approximately 10%^3,4^. Modifiable risk factors for cardiovascular disease (CVD) are often undetected or undertreated in SMI although they occur at a higher frequency^5–9^. Around half of patients with SMI are obese (body mass index (BMI) ≥ 30 kg/m^2^) or have dyslipidaemia, and the prevalence of type 2 diabetes is 3 to 5-fold higher compared to the general population^10–12^. Furthermore, patients with schizophrenia spectrum disorders have poor cardiorespiratory fitness, which has been shown to be a stronger predictor of CVD than BMI alone^13,14^.

Weight gain has a negative impact on quality of life and self-worth in patients with SMI^15,16^, which may contribute to antipsychotic non-compliance and increase the risk of relapse^17,18^. Moreover, weight gain may influence the patients’ experience of personal recovery^19^. Similarly, caregivers of patients with SMI identify weight gain as the most problematic medication-related side effect, only surpassed by sedation^20^. Metabolic comorbidity in SMI is also associated with poorer cognition ^21^. Previous studies suggest that patients with SMI are concerned about their physical health and wish to receive support from health care professionals potentially also promoting personal recovery ^22,23^. Patients with psychotic disorders often report that they receive insufficient information about their health and treatment options^24^.

Factors contributing to the high prevalence of obesity and metabolic disorders in SMI are multifaceted. SMI is associated with poor dietary habits, lower physical activity levels, and high smoking rates^25–28^. Shared risk genes with dysmetabolic disorders indicate biological links^29,30^. In addition, antipsychotic medications are associated with metabolic adverse effects including weight gain, dyslipidaemia, and risk of type 2 diabetes^31–33^. Particularly, younger and antipsychotic-naïve patients represent vulnerable individuals to these adverse effects, which appear to occur across all currently available antipsychotic agents^34^. While lifestyle interventions are recommended to treat metabolic comorbidity, insufficient adherence, and cost-effectiveness present barriers for patients with SMI^35–37^. Given contributing effects of antipsychotics to metabolic comorbidity, a switch or dose adjustment may be warranted from a physical health perspective, albeit this is not always clinically possible. Currently, no medications hold regulatory approval for management of obesity in the specific context of antipsychotics-induced weight gain. Due to the complex neurobiology and different lifestyle factors, some of the currently approved body weight loss drugs may not have the same effect in the context of antipsychotics-induced weight gain^37,38^. Due to the complexities in the underlying causes of metabolic comorbidity in SMI, interventions may need to be specifically tailored for this patient group.

Over the past two decades, while the need to enhance monitoring and management of cardiovascular risk factors has been acknowledged and highlighted in consensus guidelines, these initiatives have not produced the desired effects or been implemented in clinical practice^39^. The continued disparities in metabolic care in SMI therefore still warrant investigation of new models of care. Current Danish guidelines suggest non-specified lifestyle changes and antipsychotic switching in case of no response to lifestyle interventions, but do not include other clear recommendations regarding the management of antipsychotics-associated dysmetabolism.^40^ General practitioners (GPs) are assumed to manage metabolic comorbidity in SMI. However, these patients rarely present to the GP, or the GP may not feel comfortable addressing some of the above issues without the input of a psychiatrist; nor are GPs specialists in treating obesity. This conventional ‘silo’ working between mental and physical health constitutes an organizational barrier to access to care. For example, in a recent Canadian qualitative study, individuals with SMI were referred to an obesity clinic without expertise in mental health. Barriers identified included suboptimal care coordination and multiple appointments, challenges in establishing a good alliance with staff without experience in SMI, and lack of social support^41^. A recent European Delphi expert consensus study suggested that psychiatrists should act as the central coordinating professional in metabolic care of patients with SMI assisted as needed by other specialists (e.g. endocrinologists)^39^. Conversely, psychiatrists may experience barriers to this approach including perceived lack of training in metabolic care. Together, these issues highlight the need to adapt current models of care to facilitate collaborative interdisciplinary models and enable management of cardiometabolic health by psychiatrists.

These above-mentioned challenges may be mitigated through closer collaborations between mental and somatic caretakers and adaptation of metabolic care to the populations with SMI^42^. Most of these elements have already been implemented at the Mental Health and Metabolic Clinic at the Centre for Addiction and Mental Health (CAMH) in Toronto, Canada, but their effectiveness has not been established in rigorous methodological manner^43.^ We have designed the Meta-Care Trial as an approximated and adapted version of the CAMH approach to provide gold standard clinical evidence regarding the effectiveness of a metabolic care clinic in an independent, Danish mental health setting. Thus, the overall aim of the current randomized controlled trial (RCT) is to examine the effects of a pragmatic and individualized metabolic clinic using measurement-, evidence- and guideline-based care for antipsychotic-treated patients with SMI and significant weight gain, overweight with concomitant cardiovascular risk factors and/or obesity.

We hypothesise that 12 months of treatment in the metabolic clinic will result in 1) a greater proportion of patients achieving a weight loss of ≥5% of the initial body weight, 2) improvements of the cardiometabolic risk profile, and 3) improvements in cognition, personal recovery, and quality of life compared to standard care (outpatient clinics and/or GPs).

## Methods and analysis

The SPIRIT reporting guidelines were used for this submission.^44^

### Study population

Patients with schizophrenia spectrum disorders (International classification of diseases; ICD-10: DF2x) or bipolar disorder (ICD-10: DF30.x or DF31.x) between 18-45 years will be recruited from in- and outpatient clinics in the Mental Health Services of the Capital Region of Denmark. All patients must be legally competent and able to provide informed consent.

Inclusion criteria include: Current/ongoing treatment with antipsychotics and either a history of rapid weight gain defined as >5% body weight gain or >5 cm waist circumference increase since initiation of antipsychotic therapy; a BMI of ≥30 kg/m^2^ or a BMI >27 kg/m^2^ and concomitant cardiovascular risk factors (Hypertension defined as treatment with >1 antihypertensive drug or out-of-office/24-hour, non-invasive ambulatory blood pressure >140/90 mmHg within the previous 6 months; Dyslipidaemia defined as treatment with >1 lipid-lowering drug or elevated low-density lipoprotein (LDL) cholesterol (>3.0 mmol/l), elevated triglycerides (>1.7 mmol/l) or low high-density lipoprotein (HDL) cholesterol (<1.2 mmol/l in women and <1.0 mmol/l in men) within the previous 6 months; Sleep apnoea (ICD-10 DG473); Prediabetes or diabetes defined as HbA1c >42 mmol/mol or impaired fasting glucose > 5.6 mmol/l within the previous 6 months)^45–47^.

Reflective of the pragmatic study design, the exclusion criteria are limited to enhance generalizability of study findings to ‘real world’ clinical practice. Exclusion criteria include: clinical or laboratory evidence of comorbid medical disease not compatible with participation; unstable psychiatric disorder; severe current drug or alcohol misuse; acute suicidal risk. Application of exclusion criteria are judged by the research team constituting senior consultants in psychiatry and endocrinology.

### Study design

Patients are enrolled in an open-label, randomized controlled trial. We will recruit 84 antipsychotics-treated patients with SMI aged 18-45 years. Patients will be randomized to either the treatment arm consisting of 12 months treatment in the pragmatic metabolic clinic using individualized evidence- and guideline-based care or a standard care arm for 12 months (**Figure 1**). Allocation-ratio is 1:1. By December 2025, a total of 81 patients have been included. Data collection is expected to take place until January 2027

**Figure 1.**
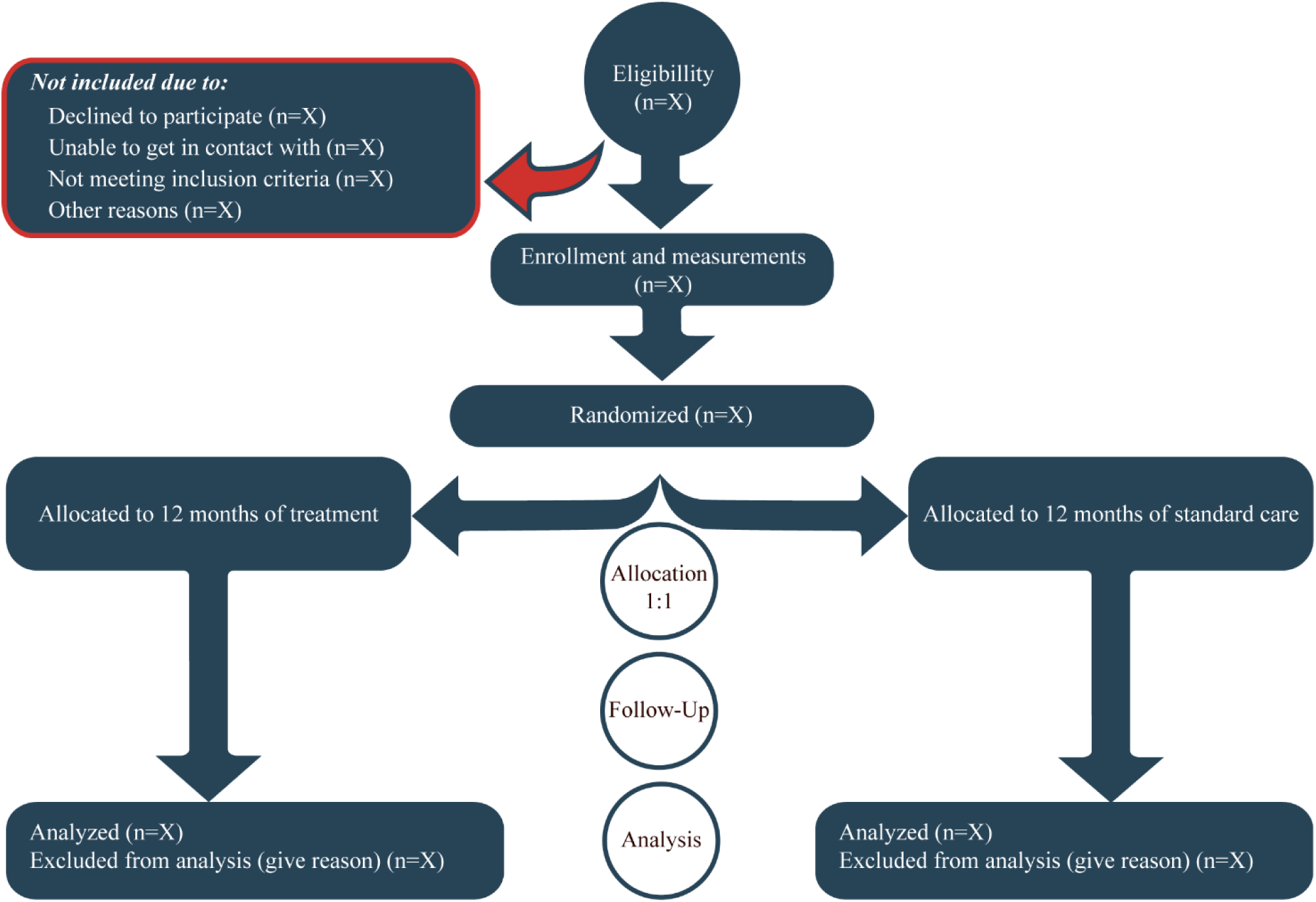
Flowchart regarding patient selection.

### Intervention

#### Meta-Care treatment arm

We will apply evidence- and measurement-based care already implemented at CAMH in Toronto, Canada^43^. The intervention includes systematic assessment of cardiometabolic measures alongside measures of cardiorespiratory fitness, body composition, cognition, personal recovery, and quality of life (**Table 1 and Figure 2**). These data are then used to drive clinical decision-making according to best practice evidence- and guideline-based treatment, patient preferences, medication profile, and physical or psychiatric comorbidities. Individualized treatment adjustments are applied when patients fail to respond adequately to the intervention or experience issues of tolerability.

**Figure 2.**
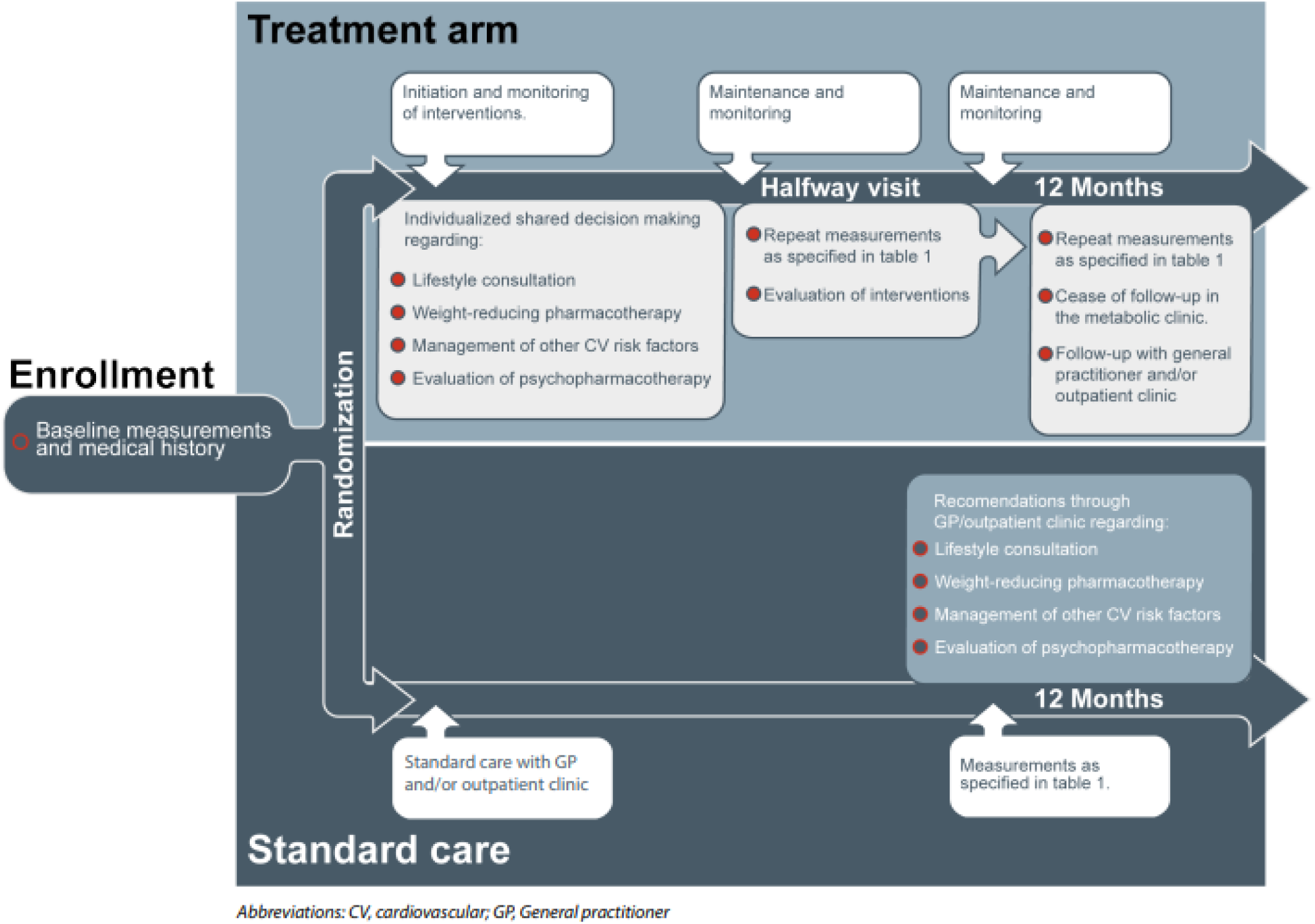
Study design with overview of the intervention and follow-up imbedded in the metabolic clinic.

**Table 1.**
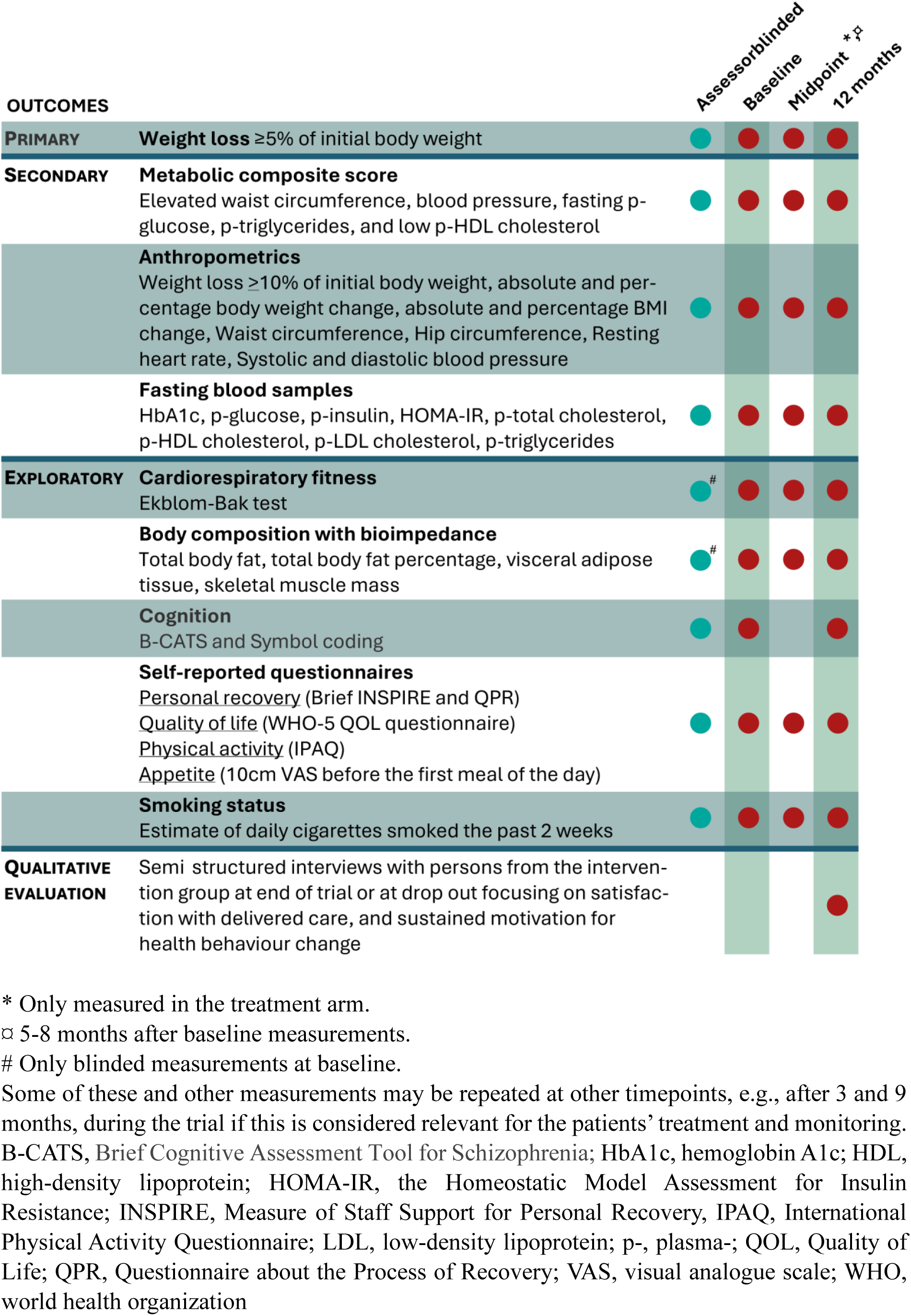
Outcomes and timepoint for measurements.

The Meta-Care treatment arm will include consultations at least once monthly with higher frequencies during titration of pharmacotherapy by medical doctors working within psychiatry (e.g., residents in psychiatry and/or specialists in general medicine) with specific metabolic training from the metabolic clinic at CAMH in Toronto, and consultations at least once monthly with an exercise physiologist in close collaboration with recognized specialists in endocrinology and psychiatry. The patients have the option to adjust duration and frequency of the consultations. The consultations will either take place as in-patient visits at Mental Health Centre Glostrup, via telephone consultations or as video consultations depending on patient preference. During the treatment period, all patients will at least once have their plan assessed at a conference with participation of the treating personnel as well as recognized specialists in endocrinology and psychiatry. Evaluation of plans at these conferences may occur multiple times during a treatment period in case of insufficient efficacy following initial assessment.

##### Treatment of cardiovascular risk factors

Management of dyslipidaemia, hypertension and type 2 diabetes including treatment with lipid-lowering pharmacotherapy, antihypertensive agents and glucose-lowering pharmacotherapy will follow Danish guidelines^46,48–50^ in close collaboration with the patients’ GP and/or recognized specialists in endocrinology.

##### Evaluation of psychotropic medication

Prescribed psychotropic medication will be evaluated in the metabolic clinic. Physicians in the metabolic clinic will not decide the patients’ psychotropic pharmacotherapy e.g. by changing prescriptions, but the metabolic clinic provides consultation and detailed recommendations to the patients’ treating psychiatrist and/or GP including considerations of psychotropic medication adjustments or switches if this is found relevant and clinically feasible to reduce the metabolic burden.

##### Pharmacological treatment of overweight and obesity

Patients have the opportunity of receiving weight-reducing pharmacotherapy. Importantly, the order of prescription of these drugs is pragmatic (i.e., not manualised) and based on an individual assessment and tailored to suit the individual patient. Thus, prescriptions will be based on patient comorbidities and shared clinical decision making and therefore does not represent a predefined, stepwise order. While the study is ongoing, updated clinical guidelines may be implemented and new drugs approved for treatment of obesity. The pragmatic study design will allow us to readily incorporate new options in the metabolic clinic, assuring patients receive updated, best practice interventions. For example, the proportion of patients receiving glucagon-like peptide 1 receptor agonists (GLP-1RAs) may increase over time given: 1) recent regulatory body approvals for management of obesity in the general population^51–53^, and 2) ongoing trials which are testing these agents in antipsychotics-treated populations^54–56^. Potential changes in practice can be incorporated in the trial as we a priori are testing the effect of the metabolic clinic as a whole and not the effect of a specific medication. Accordingly, payment of medication will follow general terms of payments for prescriptions in Denmark and will not be covered by the project. Based on patient preference, prescriptions on medication will be made by physicians in the metabolic clinic with close monitoring for known adverse events under slow titration of the included drugs or by their GP or physicians from their outpatient psychiatric treating team with recommendations from the metabolic clinic based on patient preference. Patients will only be offered pharmacotherapy with evidence to support use to mitigate antipsychotic associated weight gain (e.g., metformin) and/or pharmacotherapy licensed for treatment of overweight/obesity in Denmark (e.g., liraglutide, semaglutide, tirzepazide or naltrexone/bupropion)^57–66^.

##### Lifestyle consultations

Lifestyle consultations will be carried out with an exercise physiologist. Initially, physical activity recommendations by the Danish Health Authority and diet recommendations by the Danish Veterinary and Food Administration will be explained in relation to the patients’ own habits and behaviour^67,68^. Inclusion of relatives will be encouraged, both at consultations and by sharing content of the consultations to achieve further support. Previous exercise experience and preferences will be clarified, and exercise offers and communities designed for people with mental illness will be outlined for all patients. The consultations incorporate theory and concepts related to health behaviour change, including the Transtheoretical Model and Bandura’s theory related to self-efficacy ^69–71^. In practice, motivational factors and barriers for behaviour change will be identified with the patient, providing foundation for elucidating overall aims for participation in the intervention, and more importantly, realistic, specific, and meaningful goals in relation to diet and physical activity. This includes collaborative assessment of the patient’s current stage of change and determination of whether the previously established goals should be maintained, modified, upregulated, downregulated, or deferred, based on relevant internal or external factors. The initial consultation will be held face-to-face after randomisation. Afterwards, monthly consultations via in-patient visits, telephone or video will be held with the patient having the option to adjust duration and frequency.

#### Smoking cessation

Patients motivated for smoking cessation will be offered relevant treatment in relation to Danish guidelines for smoking cessation in persons with severe mental illness^72^. Moreover, patients will be offered referral to smoking cessation clinics for individualized counselling in the municipality where they have their address.

##### Post-study treatment

After 12 months, in-person treatment in the metabolic clinic will cease, and patients will continue follow-up with their GP and/or outpatient mental health clinic, with MD-to-MD metabolic consultations as needed. To enable assessment of sustainability, all patients will be offered follow-up with relevant measurements 12 and/or 24 months after end of participation.

#### Standard care arm

The patients who after randomisation are allocated to standard care will continue their current treatment in a psychiatric outpatient clinic and/or contact with their GP. They will receive measurements/monitoring upon enrolment and after 12 months. Following baseline measurements, the patients will be offered to have results sent to their GP and/or outpatient clinic for follow up. After 12 months, patients in standard care will be offered a lifestyle consultation similar to the initial consultation in the intervention group from an exercise physiologist and offered that a MD sends recommendations regarding the following potential post-trial interventions to the patient’s GP and/or outpatient clinic prepared in close collaboration with recognized specialists in psychiatry and endocrinology:

- Suggestions regarding relevant psychotropic medication adjustments or switches if this is found relevant and clinically feasible to reduce the metabolic burden.
- Suggestions regarding potential add-on of weight reducing pharmacotherapy.
- Suggestions regarding pharmacological treatment of other cardiovascular risk factors such as dyslipidaemia, hypertension, and type 2 diabetes.
- Suggestions regarding pharmacological treatment of patients motivated for smoking cessation and which smoking cessation clinic will be relevant for patients depending on which municipality they live in.

All patients will be offered follow-up with relevant measurements 12 and/or 24 months after end of participation.

### Measurements

Measurements and assessments during the primary study visits will be carried out during in-person visits at the Center for Neuropsychiatric Schizophrenia Research (CNSR), Mental Health Center Glostrup. After collecting informed consent by research personnel from CNSR, baseline measurements will take place. Trained scientific personnel will administer measurements and self-reported questionnaires. In case of doubts regarding individual questions from the questionnaires, the personnel will assist participants in answering them. REDCap^®^ will be used to ensure electronic data capture. Principal investigators will not have access to the final dataset. During the baseline visit, the personnel will collect data on currently used pharmacotherapy, current side effects to psychopharmacotherapy, alcohol and substance use, duration since first hospital contact related to their mental illness, family history of mental illness, socioeconomic position including current income and educational level as well as other demographics (age, sex, and ethnicity). Blood samples will be taken during fasting by trained personal under aseptic circumstances. To describe the study sample and as safety measurements, psychopathology will be evaluated using the Brief Psychiatric Rating Scale, the Clinical Global Impression scale, and the Calgary Depression Score in Schizophrenia. Patients with bipolar disorder will also have psychopathology measured using the Hamilton Depression Rating Scale and the Young Mania Rating Scale^73–77^. Level of functioning will be assessed using the Global Assessment of Functioning (GAF)^78^. Moreover, routine blood analyses, electrocardiograms and urine analyses will be carried out at least during the baseline visit. Other measurements and repeating of the baseline measures may be carried out between study visits if this is considered relevant for monitoring and treatment.

#### Study endpoints

The *primary outcome* of the RCT is the proportion of patients in the treatment group achieving a weight loss of ≥5% of the initial body weight compared to the standard care group after 12 months.

##### Rationale for choosing weight as primary outcome

From a health perspective, evidence shows that modest weight loss (i.e., ≥5% of body weight) can reduce the risk of adverse health conditions and key cardiovascular risk factors^79–81^. First-person accounts of the psychological journey of weight gain in psychosis suggest that weight gain is characterized by loss of self-worth, agency, and hope^82^.

##### Secondary outcomes include

- A metabolic composite score consisting of maximally five points (one point per composite; elevated waist circumference, elevated triglycerides, blood pressure, fasting plasma glucose, and reduced high-density lipoprotein), according to the definition and cut-off values of metabolic syndrome by the International Diabetes Federation^83,84^. *Effect measurements: differences in proportion achieving reduction of ≥1 point between groups at 12 months*.
- Proportion of patients achieving a weight loss of ≥10% of the initial body weight. *Effect measurements: differences in proportion achieving reduction of ≥10% of the initial body weight between groups at 12 months*.
- Absolute body weight change, percentage body weight change, BMI, waist circumference, hip circumference, resting heart rate and systolic/diastolic blood pressure, HbA1c, fasting plasma glucose, fasting plasma insulin, the Homeostatic Model Assessment for Insulin Resistance (HOMA-IR), fasting total cholesterol, LDL cholesterol, HDL cholesterol, and triglycerides. *Effect measurements: differences in mean changes between groups at 12 months*.
- Proportion of patients achieving a ≥50% reduction of initial fasting LDL cholesterol. *Effect measurements: differences in proportion achieving a ≥50% reduction of initial fasting LDL cholesterol levels between groups at 12 months*.

##### Exploratory outcomes include

- Cardiorespiratory fitness assessed by the submaximal Ekblom-Bak test on a mechanically braked cycle ergometer^85,86^, and body composition (total body fat, total body fat percentage, visceral adipose tissue, skeletal muscle mass) measured with a bioelectrical impedance analysis^87^. *Effect measurements: differences in mean changes between groups at 12 months*.
- Cognition measured with the Brief Cognitive Assessment Tool in Schizophrenia (B-CATS) comprised of the Trail Making Test (A and B), Letter-Number Span Record Form and Category Fluency Test (animals); and Symbol Coding ^88,89^. *Effect measurements: differences in mean changes of the individual cognitive tests between groups at 12 months*.
- Personal recovery is considered important by patients with SMI regardless of symptomatology and functioning^90^. In the current study, perceived support for personal recovery and the experience of the process of the personal recovery will be assessed with self-reporting questionnaires by means of The Brief INSPIRE Measure of Staff Support for Personal Recovery^91^ and the Questionnaire about the Process of Recovery^92^, respectively. An individual’s belief that they can achieve their desired outcomes (e.g., weight loss) is likely to be a key predictor of recovery success. *Effect measurements: differences in mean changes between groups at 12 months*.
- Quality of life measured by self-reporting questionnaire using the WHO-5 quality of life questionnaire^93^. *Effect measurements: differences in mean changes between groups at 12 months*.
- Physical activity of weekly self-perceived volumes of different intensities and average daily sedentary hours measured by the self-reporting questionnaire, the International Physical Activity Questionnaire^94^. *Effect measurements: differences in mean changes between groups at 12 months*.
- Appetite measured by self-reporting, using a 10cm visual analogue scale before the first meal of the day^95^. *Effect measurements: differences in mean changes between groups at 12 months*.
- Smoking cessation amongst persons who are smoking at baseline measured as self-reported cessation for the past 7 days after 12 months and average total daily use of cigarettes for the past week. *Effect measurements: differences in mean changes of reported daily use of cigarettes between groups at 12 months and proportion of persons smoking at baseline with smoking cessation*.

#### Qualitative evaluation

Post-intervention semi-structured interviews will be conducted with a purposefully sampled subgroup of participants. The sampling aims to ensure rich data and maximum variation in terms of sex, age, and diagnosis. The interviews will explore satisfaction with the delivered care, and motivation for health behaviour change. Moreover, all dropouts (i.e., convenience sampling) will be invited to participate in an interview regarding their reasons for dropping out. Adequate sample size for the qualitative evaluation will be guided by information power (also denoted saturation)^96,97^.

### Randomization and blinding

Eligible patients will be randomized in a 1:1 ratio stratified for sex using random number tables with random block sizes varying between 2, 4, or 6. A randomisation list generator with a blinded random seed and defined block sizes were used to program the randomization sequence. The sequence will be uploaded to REDCap where the randomisation will be carried out. Staff from CNSR not involved in data collection will program the randomization sequence. Randomizations occur after baseline assessments on the first examination day to ensure blinding of involved staff before allocation to either the treatment group or the standard care group. Moreover, research personnel conducting the data collection of primary and secondary outcomes as well as cognition will be blinded to patients group allocation at halfway and 12 months visits (see **Table 1**). Participants will be reminded prior to each follow-up assessment not to reveal their group allocation. Blinding of patients and the treatment staff (i.e. medical doctors and the exercise physiologist) will not be possible following randomization after baseline measurements due to the nature of the trial.

### Sample size and power calculation

The sample size calculation is based on unpublished results from CAMH in Toronto, Canada, and similar power calculations testing dichotomous variables with differences of proportions in two groups over time^43^. Presumed that ≥5% weight loss after 12 months is achieved by 45% in the intervention group, which is deemed an effect size a priori, compared with 15% in the control group, the null hypothesis that the proportion achieving ≥5% weight loss after 12 months in each group is equal, can be rejected with a probability of 0.8 (ß) if 70 patients are included. The probability of a type I error associated with this null hypothesis test is 0.05 (α). To account for 15% attrition rates observed at CAMH, the present study aims to include a total of 84 patients.

### Analyses plan

Conventional statistical tests (e.g., t tests, chi2) and standardized effect sizes will be used to describe the profile of the sample and investigate group differences at baseline. We will use logistic regression to estimate the effect of the intervention on our primary (≥5% reduction in weight) and secondary dichotomous outcomes while controlling for the stratification variable sex and baseline weight for our primary/secondary model, respectively, which is expected to be associated with the outcome thus contributing for improved power. The group differences at 12 months will be reported in terms of odds ratios and model estimated prevalence along with their precision (95% confidence intervals) and declared significant if p-value <0.05. A sensitivity analysis will be conducted adjusting for variables that show larger than medium effect size between groups at baseline, where effect sizes are measured by Cohen’s d or Cramer’s V for continuous and categorical variables, respectively. We will also conduct a sensitivity analysis using multiple imputation to account for dropouts and individuals lost during follow up. The imputation model will include baseline information mentioned as covariates above, and possibly others that may be found to be associated with weight at 12 months, like baseline weight. Additionally, we will report a comparison between completers and non-completers regarding their baseline characteristics to inform about possible reasons for dropout and investigate the potential for biases due to missing data. With regard to our continuous secondary outcomes, an analysis of covariance (ANCOVA) model will be used to compare groups at 12 months controlling for baseline continuous secondary variables and the stratification variable sex. Since the ANCOVA model can only use complete cases, multiple imputation will be used to account for missingness. Mean difference at 12 months and 95% confidence intervals will be reported. Model diagnostic analysis will be conducted to access assumptions (normality of residuals, homogeneity of variance, influential data points and linearity of individual continuous variables), and violation of assumptions will be addressed with sensitivity analyses (transformation or generalized linear model for non-normal residuals, refitting the models without influential points, generalized least square for non-constant variance, generalized additive models for evidence of non-linearity of individual baseline continuous variables). Adverse events between groups will be analysed using Fisher’s exact test.

In exploratory analyses, sex will be added as a moderator by specifying the interaction between sex and treatment group in the logistic and ANCOVA models. In order to ensure all sex-relevant information is conveyed and considering that the moderation analysis is secondary without the study being powered for it, we will report estimates of treatment effect by sex regardless of whether sex is found to play a statistically significant role as a moderator. The data management and analyses will be conducted with SAS version 9.4 (SAS institute Inc., NC, USA) and package lme4 in R.

Qualitative data will be analysed by means of inductive-deductive thematic analysis informed by the COM-B model identifying capability, opportunity, and motivation as key factors which need to change in order for a behaviour change intervention to be effective^97^. Adequate sample size for the qualitative evaluation will be guided by information power (also denoted saturation)^98^.

### Patient and public involvement

We will engage with stakeholders (e.g., patients working as recovery mentors, patient organizations, clinicians, participating patients via post-participation interviews and managers/decision makers) throughout the entire research process on development of recruitment material, study information, consent forms and optimizations of the interventions.

### Ethics and dissemination

#### Safety

There are few potential risks related to participating in the study, which are primarily related to the known side effects of the pharmacotherapy patients may be offered^46,48,49,57–66^. As mentioned, we will apply relevant psychometrics to estimate deterioration of mental health leading to relevant safety procedures. Adverse events are any adverse or unintentional reactions as well as any symptoms which might occur during the trial period regardless of a potential association with the study participation. The adverse events including severe adverse events will be registered in case report files of the individual participants via REDCap®. In addition to collection of adverse events during study visits, information regarding healthcare contacts and inpatient admissions in somatic and psychiatric emergency departments will be registered from the participants’ medical record regardless of which group they are allocated to.

#### Ethical considerations

Ethical permission for the trial has been granted by the Regional Ethics Committee (ID: H-23045618) and approval has been granted by the Data Protection Agency in the Capital Region of Copenhagen (ID: p-2023-14342). Investigators will respect the law on handling of personal data. This study will be performed according to the Declaration of Helsinki (2008) and the International Conference on Harmonisation – Good Clinical Practice (ICH-GCP). Since the Danish Medicines Agency do not consider this a trial with medicinal products (correspondence case number 2023031879), application for permission will not be applied to the Danish Medicines Agency. The trial is registered at ClinicalTrials.gov and will conform to GCP guidelines, albeit the project is not monitored by the GCP unit in the capital region of Copenhagen since the trial is not categorized as a trial with medicinal products. All participants in the study are covered by the patient injury insurance and by the general insurance for patients treated in public hospitals (Danish: Patienterstatningsordningen).

Any changes to the protocol will be documented in protocol amendments, which must be approved by the Ethics Committee for the Capital Region of Denmark and reported when the study is disseminated. Study sites and recruitment status will be available at clinicaltrials.gov and all participants will receive best practice.

As underlined in previous paragraphs, the anticipated risks of serious adverse events in relation to participation is considered low. In contrast, potential substantial benefits of the intervention on factors such as weight loss, reduction of cardiovascular risk factors, CVD, T2D and other non-cardiovascular domains such as cognition and self-worth are anticipated. Therefore, to the best of our judgement, the benefits of the study outweigh the risk of adverse events.

#### Clinical significance

This will be the first RCT to investigate the effectiveness of a pragmatic and individualised metabolic clinic for SMI patients receiving antipsychotic pharmacotherapy with associated weight gain and metabolic disorders versus standard care in a regional mental health organization. The combined quantitative and qualitative data will provide detailed insights into the active ingredients related to effectiveness, which are of pivotal importance for ensuring sustainability and implementation of the intervention beyond the end of the trial. Person-centred indicators of the success of the metabolic clinic will be captured by scales of personal recovery and health care support. If the results are positive, the Meta-Care trial can easily be implemented within the existing structure of psychiatric practice in Denmark, thereby beginning to break down historical silos of care.

In longer term perspective, we foresee: **1)** Translation of evidence-based knowledge into clinical treatment of patients with SMI and comorbid modifiable cardiovascular risk factors; **2)** increased medical knowledge and improved metabolic management by psychiatrists; **3)** enablement of interdisciplinary collaborations and improved collaborations between GPs and psychiatrists, and **4)** establishment of well-defined standards of metabolic monitoring and care.

Beyond physical health improvements, implementation of a metabolic clinic has the potential of providing better basis for patients’ desired active involvement in decisions about their health care and is in accordance with The Lancet Psychiatry Commission’s recommendations for protecting physical health in patients with SMI^99^. The clinic will promote integration of historically separated services, facilitating adaptions in service structure, delivery, and culture instead of an isolated focus on individual behavioural change or a single weight loss medication.

#### Dissemination

Positive, negative, or inconclusive results will be published in international scientific peer-reviewed journals and presented at national and international conferences. No data will be sent abroad unless requested for individual patient data analyses. In this case, pseudo anonymized data will be shared after signing a data access agreement.

## Data Availability

No data will be sent abroad unless requested for individual patient data analyses. In this case, pseudo anonymized data will be shared after signing a data access agreement.

## Contributions

BHE and MKH were overall responsible for designing and conducting the trial. BHE is the sponsor while BHE and MKH are principal investigators of the study. BHE and MKE have conceived the idea and promoted the concept of the study with contributions from GHM, SMA, VS, MS, MES, DS, TV, JM and CKL. CKL, GHM, SMA, MKH and BHE were involved in designing the cognitive and psychopathological part of the study. JM was responsible for designing the qualitative evaluation. BHE, MKH, GHM, SMA, and MS were primarily involved in developing the statistical analysis plan. GHM and SMA contributed equally to this paper and share first authorship. BHE and MKH share senior authorship. All authors have made substantial contributions to the conception and design of the study as well as the drafting and critical revision of the manuscript. All authors have read and approved the final version of the manuscript and agree to be accountable for all aspects of the work, ensuring that questions related to the accuracy or integrity of the work are appropriately investigated and resolved.

## Competing interest statement

BHE is part of the Advisory Board of Boehringer Ingelheim, Lundbeck Pharma, and Orion Pharma; and has received lecture fees from Boehringer Ingelheim, Otsuka Pharma Scandinavia AB, and Lundbeck Pharma.

DS is on a Quality Assurance Committee for Viatris

SMA has received lecture fees from HLS therapeutics and Boehringer Ingelheim, Canada.

TV has served on scientific advisory panels, been part of speaker’s bureaus, served as a consultant to and/or received research support from Amgen, AstraZeneca, Boehringer Ingelheim, Eli Lilly, Gilead, GSK, Mundipharma, MSD/Merck, Novo Nordisk, Regor, Roche, Sanofi, Sun Pharmaceuticals and Zealand Pharma

The remaining authors have no competing interests to declare.

## References

1. Nielsen RE, Uggerby AS, Jensen SOW, McGrath JJ. Increasing mortality gap for patients diagnosed with schizophrenia over the last three decades--a Danish nationwide study from 1980 to 2010. Schizophr Res. 2013 May;146(1–3):22–7.

2. Lindstrӧm C, Siersma V, Kriegbaum M, Grauers Willadsen T, Bakkedal C, Brodersen JB, et al. Time trends in mortality for people with severe mental illness in Denmark 2000-2018. Nord J Psychiatry. 2025 Jan;79(1):79–85.

3. Sher L, Kahn RS. Suicide in Schizophrenia: An Educational Overview. Medicina (Kaunas). 2019 Jul;55(7).

4. Halstead S, Cao C, Høgnason Mohr G, Ebdrup BH, Pillinger T, McCutcheon RA, et al. Prevalence of multimorbidity in people with and without severe mental illness: a systematic review and meta-analysis. Lancet Psychiatry [Internet]. 2024;11:431–42. Available from: https://www.sciencedirect.com/science/article/pii/S2215036624000919

5. Nasrallah HA, Meyer JM, Goff DC, McEvoy JP, Davis SM, Stroup TS, et al. Low rates of treatment for hypertension, dyslipidemia and diabetes in schizophrenia: data from the CATIE schizophrenia trial sample at baseline. Schizophr Res. 2006 Sep;86(1–3):15–22.

6. Mitchell AJ, Lord O, Malone D. Differences in the prescribing of medication for physical disorders in individuals with v. without mental illness: meta-analysis. Br J Psychiatry. 2012 Dec;201(6):435–43.

7. Mitchell AJ, Delaffon V, Vancampfort D, Correll CU, De Hert M. Guideline concordant monitoring of metabolic risk in people treated with antipsychotic medication: systematic review and meta-analysis of screening practices. Psychol Med. 2012 Jan;42(1):125–47.

8. Bakkedal C, Persson F, Christensen MB, Kriegbaum M, Mohr GH, Andersen JS, et al. The development of type 2 diabetes management in people with severe mental illness in the Capital Region of Denmark from 2001 to 2015. Acta Psychiatr Scand. 2024 Mar;149(3):219–33.

9. Høgnason Mohr G, Barcella CA, Grand MK, Kriegbaum M, Siersma V, Hahn MK, et al. Management of dyslipidaemia in individuals with severe mental illness: a population-based study in the Greater Copenhagen Area. Ther Adv Psychopharmacol. 2023;13:20451253231211576.

10. Dixon L, Weiden P, Delahanty J, Goldberg R, Postrado L, Lucksted A, et al. Prevalence and correlates of diabetes in national schizophrenia samples. Schizophr Bull. 2000;26(4):903–12.

11. Mukherjee S, Decina P, Bocola V, Saraceni F, Scapicchio PL. Diabetes mellitus in schizophrenic patients. Compr Psychiatry. 1996;37(1):68–73.

12. De Hert M, van Winkel R, Van Eyck D, Hanssens L, Wampers M, Scheen A, et al. Prevalence of diabetes, metabolic syndrome and metabolic abnormalities in schizophrenia over the course of the illness: a cross-sectional study. Clin Pract Epidemiol Ment Health. 2006 Jun;2:14.

13. Weeldreyer NR, De Guzman JC, Paterson C, Allen JD, Gaesser GA, Angadi SS. Cardiorespiratory fitness, body mass index and mortality: a systematic review and meta-analysis. Br J Sports Med. 2025 Feb;59:339–46.

14. Scheewe TW, van Haren NEM, Sarkisyan G, Schnack HG, Brouwer RM, de Glint M, et al. Exercise therapy, cardiorespiratory fitness and their effect on brain volumes: A randomised controlled trial in patients with schizophrenia and healthy controls. European Neuropsychopharmacology. 2013;23(7):675–85.

15. Faulkner G, Cohn T, Remington G. Interventions to reduce weight gain in schizophrenia. Cochrane Database Syst Rev. 2007 Jan;(1):CD005148.

16. Fakhoury WK, Wright D, Wallace M. Prevalence and extent of distress of adverse effects of antipsychotics among callers to a United Kingdom National Mental Health Helpline. Int Clin Psychopharmacol. 2001 May;16(3):153–62.

17. Velligan DI, Weiden PJ, Sajatovic M, Scott J, Carpenter D, Ross R, et al. The expert consensus guideline series: adherence problems in patients with serious and persistent mental illness. J Clin Psychiatry. 2009;70 Suppl 4:1–8.

18. De R, Smith ECC, Navagnanavel J, Au E, Maksyutynska K, Papoulias M, et al. The impact of weight gain on antipsychotic nonadherence or discontinuation: A systematic review and meta-analysis. Acta Psychiatr Scand. 2025 Feb;151(2):109–26.

19. Leamy M, Bird V, Le Boutillier C, Williams J, Slade M. Conceptual framework for personal recovery in mental health: systematic review and narrative synthesis. Br J Psychiatry. 2011 Dec;199:445–52.

20. Angermeyer MC, Matschinger H. [Neuroleptics and quality of life. A patient survey]. Psychiatr Prax. 2000 Mar;27(2):64–8.

21. Li C, Zhan G, Rao S, Zhang H. Metabolic syndrome and its factors affect cognitive function in chronic schizophrenia complicated by metabolic syndrome. J Nerv Ment Dis. 2014 Apr;202(4):313–8.

22. Schnor H, Linderoth S, Midtgaard J. Patient and Mental Health Care Professionals’ Perspectives on Health Promotion in Psychiatric Clinical Practice: A Focus Group Study. Issues Ment Health Nurs. 2021 Sep;42(9):870–9.

23. Farholm A, Sørensen M. Motivation for physical activity and exercise in severe mental illness: A systematic review of intervention studies. Int J Ment Health Nurs. 2016 Jun;25(3):194–205.

24. Haugom EW, Stensrud B, Beston G, Ruud T, Landheim AS. Experiences of shared decision making among patients with psychotic disorders in Norway: a qualitative study. BMC Psychiatry. 2022 Mar;22(1):192.

25. Bueno-Antequera J, Oviedo-Caro MÁ, Munguía-Izquierdo D. Sedentary behaviour, physical activity, cardiorespiratory fitness and cardiometabolic risk in psychosis: The PsychiActive project. Schizophr Res. 2018;195:142–8.

26. Vancampfort D, Probst M, Scheewe T, De Herdt A, Sweers K, Knapen J, et al. Relationships between physical fitness, physical activity, smoking and metabolic and mental health parameters in people with schizophrenia. Psychiatry Res. 2013;207(1–2):25–32.

27. Jakobsen AS, Speyer H, Nørgaard HCB, Karlsen M, Hjorthøj C, Krogh J, et al. Dietary patterns and physical activity in people with schizophrenia and increased waist circumference. Schizophr Res. 2018;199:109–15.

28. Andersen E, Holmen TL, Egeland J, Martinsen EW, Bigseth TT, Bang-Kittilsen G, et al. Physical activity pattern and cardiorespiratory fitness in individuals with schizophrenia compared with a population-based sample. Schizophr Res. 2018;201:98–104.

29. Amare AT, Schubert KO, Klingler-Hoffmann M, Cohen-Woods S, Baune BT. The genetic overlap between mood disorders and cardiometabolic diseases: a systematic review of genome wide and candidate gene studies. Transl Psychiatry. 2017 Jan;7:e1007.

30. Andreassen OA, Djurovic S, Thompson WK, Schork AJ, Kendler KS, O’Donovan MC, et al. Improved detection of common variants associated with schizophrenia by leveraging pleiotropy with cardiovascular-disease risk factors. Am J Hum Genet. 2013 Feb;92:197–209.

31. Rajkumar AP, Horsdal HT, Wimberley T, Cohen D, Mors O, Børglum AD, et al. Endogenous and Antipsychotic-Related Risks for Diabetes Mellitus in Young People With Schizophrenia: A Danish Population-Based Cohort Study. Am J Psychiatry. 2017 Jul;174(7):686–94.

32. De Hert M, Detraux J, van Winkel R, Yu W, Correll CU. Metabolic and cardiovascular adverse effects associated with antipsychotic drugs. Nat Rev Endocrinol. 2011 Oct;8(2):114–26.

33. Ballon JS, Pajvani U, Freyberg Z, Leibel RL, Lieberman JA. Molecular pathophysiology of metabolic effects of antipsychotic medications. Trends Endocrinol Metab. 2014 Nov;25(11):593–600.

34. Zipursky RB, Gu H, Green AI, Perkins DO, Tohen MF, McEvoy JP, et al. Course and predictors of weight gain in people with first-episode psychosis treated with olanzapine or haloperidol. Br J Psychiatry. 2005 Dec;187:537–43.

35. Holt RIG, Gossage-Worrall R, Hind D, Bradburn MJ, McCrone P, Morris T, et al. Structured lifestyle education for people with schizophrenia, schizoaffective disorder and first-episode psychosis (STEPWISE): Randomised controlled trial. British Journal of Psychiatry. 2019;214(2):63–73.

36. Speyer H, Christian Brix Nørgaard H, Birk M, Karlsen M, Storch Jakobsen A, Pedersen K, et al. The CHANGE trial: No superiority of lifestyle coaching plus care coordination plus treatment as usual compared to treatment as usual alone in reducing risk of cardiovascular disease in adults with schizophrenia spectrum disorders and abdominal obesity. World Psychiatry. 2016;15(2):155–65.

37. Obesity Canada. The Role of Mental Health in Obesity Management [Internet]. Canadian Adult Obesity Clinical Practice Guidelines. 2020. p. (webpage). Available from: https://obesitycanada.ca/wp-content/uploads/2021/05/7-The-Role-of-Mental-Health-v4-with-links.pdf

38. Joffe G, Takala P, Tchoukhine E, Hakko H, Raidma M, Putkonen H, et al. Orlistat in clozapine-or olanzapine-treated patients with overweight or obesity: a 16-week randomized, double-blind, placebo-controlled trial. J Clin Psychiatry. 2008 May;69:706–11.

39. Galderisi S, De Hert M, Del Prato S, Fagiolini A, Gorwood P, Leucht S, et al. Identification and management of cardiometabolic risk in subjects with schizophrenia spectrum disorders: A Delphi expert consensus study. European Psychiatry. 2021;64(1).

40. Medicinrådet. Baggrund for Medicinrådets behandlingsvejledning vedrørende antipsykotika til behandling af psykotiske tilstande hos voksne. Medicinrådet. 2020.

41. Melamed OC, Fernando I, Soklaridis S, Hahn MK, Frcp C, Lemessurier KW, et al. Understanding Engagement with a Physical Health Service : A Qualitative Study of Patients with Severe Mental Illness. The Canadian Journal of Psychiatry. 2019;64(12):872–80.

42. Druss BG, von Esenwein SA, Glick GE, Deubler E, Lally C, Ward MC, et al. Randomized Trial of an Integrated Behavioral Health Home: The Health Outcomes Management and Evaluation (HOME) Study. Am J Psychiatry. 2017 Mar;174(3):246–55.

43. The Centre for Addiction and Mental Health. Mental Health and Metabolism Clinic (CAMH) [Internet]. Mental Health and Metabolism Clinic. 2025 [cited 2025 Apr 15]. p. (webpage). Available from: https://www.camh.ca/en/patients-and-families/programs-and-services/mental-health-metabolism-clinic

44. Chan AW, Tetzlaff JM, Gøtzsche PC, Altman DG, Mann H, Berlin JA, et al. SPIRIT 2013 explanation and elaboration: guidance for protocols of clinical trials. BMJ. 2013 Jan;346:e7586.

45. Alberti KGMM, Zimmet P, Shaw J. Metabolic syndrome - A new world-wide definition. A consensus statement from the International Diabetes Federation. Diabetic Medicine. 2006;23(5):469–80.

46. Dyslipidæmi [Internet]. Dansk Cardiologisk Selskab (DCS). 2024 [cited 2025 Apr 15]. Available from: https://nbv.cardio.dk/dyslipidaemi

47. Alberti KGMM, Eckel RH, Grundy SM, Zimmet PZ, Cleeman JI, Donato KA, et al. Harmonizing the metabolic syndrome: A joint interim statement of the international diabetes federation task force on epidemiology and prevention; National heart, lung, and blood institute; American heart association; World heart federation; International . Circulation. 2009;120(16):1640–5.

48. Arteriel Hypertension [Internet]. Dansk Cardiologisk Selskab (DCS). 2024 [cited 2025 Apr 15]. Available from: https://nbv.cardio.dk/hypertension

49. Type 2 diabetes [Internet]. Dansk Endokrinologisk Selskab. 2024. Available from: https://endocrinology.dk/nbv/diabetes-melitus/behandling-og-kontrol-af-type-2-diabetes/

50. Forebyggelse af hjertesygdom [Internet]. Dansk Cardiologisk Selskab (DCS). 2024 [cited 2025 Apr 15]. Available from: https://nbv.cardio.dk/forebyggelse

51. U.S Food and Drug adminstration. FDA Approves new drug treatments for chronic weight management. [Internet]. FDA Drug Safety Communication. 2022. Available from: https://www.fda.gov/news-events/press-announcements/fda-approves-new-drug-treatment-chronic-weight-management-first-2014

52. Wilding JPH, Batterham RL, Calanna S, Davies M, Van Gaal LF, Lingvay I, et al. Once-Weekly Semaglutide in Adults with Overweight or Obesity. N Engl J Med. 2021 Mar;384(11):989–1002.

53. Sundhedsstyrelsen. Wegovy (semaglutid) Præparatanmeldelse [Internet]. 2022 [cited 2023 Mar 13]. Available from: https://www.sst.dk/-/media/Viden/Laegemidler/Praeparatanmeldelser/Wegovy/Praeparatanmeldelse-Wegovy_FINAL_161222.ashx?sc_lang=da&hash=46C929E8F36F50B0F3793D750FE9556B

54. Sass MR, Danielsen AA, Köhler-Forsberg O, Storgaard H, Knop FK, Nielsen MØ, et al. Effect of the GLP-1 receptor agonist semaglutide on metabolic disturbances in clozapine-treated or olanzapine-treated patients with a schizophrenia spectrum disorder: study protocol of a placebo-controlled, randomised clinical trial (SemaPsychiatry). BMJ Open. 2023 Jan;13:e068652.

55. Hahn MK. Semaglutide in Comorbid Schizophrenia Spectrum Disorder and Obesity (Sema) [Internet]. 2022 [cited 2023 Mar 13]. Available from: https://clinicaltrials.gov/ct2/show/NCT05333003

56. Frystyk J. Home-based Intervention With Semaglutide Treatment Of Neuroleptica-Related Prediabetes (HISTORI) [Internet]. 2022 [cited 2023 Mar 13]. Available from: https://clinicaltrials.gov/ct2/show/NCT05193578

57. Siskind DJ, Leung J, Russell AW, Wysoczanski D, Kisely S. Metformin for Clozapine Associated Obesity: A Systematic Review and Meta-Analysis. PLoS One. 2016;11(6):e0156208.

58. Praharaj SK, Jana AK, Goyal N, Sinha VK. Metformin for olanzapine-induced weight gain: a systematic review and meta-analysis. Br J Clin Pharmacol. 2011 Mar;71(3):377–82.

59. Fitzgerald I, O’Connell J, Keating D, Hynes C, McWilliams S, Crowley EK. Metformin in the management of antipsychotic-induced weight gain in adults with psychosis: development of the first evidence-based guideline using GRADE methodology. Evid Based Ment Health. 2022;25(1):15–22.

60. Saxenda®, produktinformation [Internet]. Pro.medicin.dk. 2025 [cited 2025 Apr 13]. Available from: https://pro.medicin.dk/Medicin/Praeparater/7837

61. Pro.medicin.dk. Semaglutid, produktinformation [Internet]. 2023 [cited 2023 Mar 13]. Available from: https://pro.medicin.dk/Medicin/Praeparater/10247

62. Okuyama Y, Oya K, Matsunaga S, Kishi T, Iwata N. Efficacy and tolerability of topiramate-augmentation therapy for schizophrenia: a systematic review and meta-analysis of randomized controlled trials. Neuropsychiatr Dis Treat. 2016;12:3221–36.

63. Hahn MK, Cohn T, Teo C, Remington G. Topiramate in schizophrenia: a review of effects on psychopathology and metabolic parameters. Clin Schizophr Relat Psychoses. 2013 Jan;6(4):186–96.

64. Goh KK, Chen CH, Lu ML. Topiramate mitigates weight gain in antipsychotic-treated patients with schizophrenia: meta-analysis of randomised controlled trials. Int J Psychiatry Clin Pract. 2019 Mar;23(1):14–32.

65. Pro.medicin.dk. Mysimba, produktinformation [Internet]. 2022 [cited 2023 Mar 13]. Available from: https://pro.medicin.dk/Medicin/Praeparater/8479

66. Mounjaro, produktinformation [Internet]. Pro.medicin.dk. 2025. Available from: https://pro.medicin.dk/Medicin/Praeparater/10641

67. Bevæg dig på din måde - helst 30 minutter om dagen [Internet]. Sundhedsstyrelsen. [cited 2025 Apr 13]. Available from: https://www.sst.dk/da/30min

68. Kostråd til dig [Internet]. Fødevarestyrelsen. [cited 2025 Apr 13]. Available from: https://foedevarestyrelsen.dk/kost-og-foedevarer/alt-om-mad/de-officielle-kostraad/kostraad-til-dig

69. Bandura A. Self-efficacy: toward a unifying theory of behavioral change. Psychol Rev. 1977 Mar;84(2):191–215.

70. Davis R, Campbell R, Hildon Z, Hobbs L, Michie S. Theories of behaviour and behaviour change across the social and behavioural sciences: a scoping review. Health Psychol Rev. 2015;9(3):323–44.

71. Prochaska JO, Velicer WF. The transtheoretical model of health behavior change. Am J Health Promot. 1997;12(1):38–48.

72. Rygning og rygestop blandt patienter med psykiske lidelser – en guide til praktiserende læger og læger i hospitals- og distriktspsykiatrien [Internet]. Sundhedsstyrelsen. 2022 [cited 2025 Apr 13]. Available from: https://www.sst.dk/-/media/Udgivelser/2022/Rygestop-og-psykiatri/RYGNING_PSYKISKE_LIDELSER_LAEGER.ashx?sc_lang=da&hash=8608081F8FC6174FEB05F9EC07D551E6

73. Young RC, Biggs JT, Ziegler VE, Meyer DA. A rating scale for mania: Reliability, validity and sensitivity. British Journal of Psychiatry. 1978;133(11):429–35.

74. Hamilton M. A rating scale for depression. J Neurol Neurosurg Psychiatry. 1960 Feb;23(1):56–62.

75. Overall JE, Gorham DR. The Brief Psychiatric Rating Scale. Psychol Rep. 1962;10.

76. Busner J, Targum SD. The clinical global impressions scale: applying a research tool in clinical practice. Psychiatry (Edgmont). 2007 Jul;4:28–37.

77. Addington D, Addington J, Maticka-Tyndale E. Assessing depression in schizophrenia: the Calgary Depression Scale. Br J Psychiatry Suppl. 1993 Dec;(22):39–44.

78. Hall RC. Global assessment of functioning. A modified scale. Psychosomatics. 1995;36:267–75.

79. Avenell A, Broom J, Brown TJ, Poobalan A, Aucott L, Stearns SC, et al. Systematic review of the long-term effects and economic consequences of treatments for obesity and implications for health improvement. Health Technol Assess. 2004 May;8(21):iii–iv, 1–182.

80. Knowler WC, Fowler SE, Hamman RF, Christophi CA, Hoffman HJ, Brenneman AT, et al. 10-year follow-up of diabetes incidence and weight loss in the Diabetes Prevention Program Outcomes Study. Lancet. 2009 Nov;374(9702):1677–86.

81. Tuomilehto J, Lindström J, Eriksson JG, Valle TT, Hämäläinen H, Ilanne-Parikka P, et al. Prevention of type 2 diabetes mellitus by changes in lifestyle among subjects with impaired glucose tolerance. N Engl J Med. 2001 May;344(18):1343–50.

82. Waite F, Langman A, Mulhall S, Glogowska M, Hartmann-Boyce J, Aveyard P, et al. The psychological journey of weight gain in psychosis. Psychol Psychother. 2022 Feb;

83. Alberti KGMM, Zimmet P, Shaw J. Metabolic syndrome - A new world-wide definition. A consensus statement from the International Diabetes Federation. Diabetic Medicine. 2006;23(5):469–80.

84. Alberti KGMM, Eckel RH, Grundy SM, Zimmet PZ, Cleeman JI, Donato KA, et al. Harmonizing the metabolic syndrome: A joint interim statement of the international diabetes federation task force on epidemiology and prevention; National heart, lung, and blood institute; American heart association; World heart federation; International . Circulation. 2009;120(16):1640–5.

85. Björkman F, Ekblom-Bak E, Ekblom Ö, Ekblom B. Validity of the revised Ekblom Bak cycle ergometer test in adults. Eur J Appl Physiol. 2016;116(9):1627–38.

86. Ekblom-Bak E, Björkman F, Hellenius ML, Ekblom B. A new submaximal cycle ergometer test for prediction of VO2max. Scand J Med Sci Sports. 2014;24(2):319–26.

87. Khalil SF, Mohktar MS, Ibrahim F. The theory and fundamentals of bioimpedance analysis in clinical status monitoring and diagnosis of diseases. Sensors (Basel). 2014 Jun;14(6):10895–928.

88. Hurford IM, Ventura J, Marder SR, Reise SP, Bilder RM. A 10-minute measure of global cognition: Validation of the Brief Cognitive Assessment Tool for Schizophrenia (B-CATS). Schizophr Res. 2018 May 1;195:327–33.

89. Keefe RSE, Goldberg TE, Harvey PD, Gold JM, Poe MP, Coughenour L. The Brief Assessment of Cognition in Schizophrenia: reliability, sensitivity, and comparison with a standard neurocognitive battery. Schizophr Res. 2004 Jun;68(2–3):283–97.

90. Skar-Fröding R, Clausen HK, Šaltytė Benth J, Ruud T, Slade M, Sverdvik Heiervang K. The Importance of Personal Recovery and Perceived Recovery Support Among Service Users With Psychosis. Psychiatr Serv. 2021 Jun;72(6):661–8.

91. Williams J, Leamy M, Bird V, Le Boutillier C, Norton S, Pesola F, et al. Development and evaluation of the INSPIRE measure of staff support for personal recovery. Soc Psychiatry Psychiatr Epidemiol. 2015 May;50(5):777–86.

92. Neil ST, Kilbride M, Pitt L, Nothard S, Welford M, Sellwood W, et al. The questionnaire about the process of recovery (QPR): A measurement tool developed in collaboration with service users. Psychosis. 2009 Aug;1(2):145–55.

93. World Health Organization. Regional Office for Europe. WELLBEING MEASURES IN PRIMARY HEALTH CARE/ THE DEPCARE PROJECT Report on a WHO Meeting. Stockholm; 1998.

94. Hagströmer M, Oja P, Sjöström M. The International Physical Activity Questionnaire (IPAQ): a study of concurrent and construct validity. Public Health Nutr. 2006 Sep;9:755–62.

95. Flint A, Raben A, Blundell JE, Astrup A. Reproducibility, power and validity of visual analogue scales in assessment of appetite sensations in single test meal studies [Internet]. Vol. 24, International Journal of Obesity. 2000. Available from: www.nature.com/ijo

96. Malterud K, Siersma VD, Guassora AD. Sample Size in Qualitative Interview Studies: Guided by Information Power. Qual Health Res. 2016 Nov;26(13):1753–60. 6

97. Michie S, van Stralen MM, West R. The behaviour change wheel: a new method for characterising and designing behaviour change interventions. Implementation Science. 2011 Apr;6:42.

98. Malterud K, Siersma VD, Guassora AD. Sample Size in Qualitative Interview Studies: Guided by Information Power. Qual Health Res. 2016 Nov;26(13):1753–60.

99. Firth J, Siddiqi N, Koyanagi A, Siskind D, Rosenbaum S, Galletly C, et al. The Lancet Psychiatry Commission: a blueprint for protecting physical health in people with mental illness. Lancet Psychiatry. 2019 Aug;6(8):675–712.

